# Upper Extremity Muscle Recovery Profiles with Functional Electrical Stimulation Therapy in Chronic Spinal Cord Injury

**DOI:** 10.1101/2025.05.27.25328361

**Authors:** Gustavo Balbinot, Guijin Li, Alexandra Chen, Parvin Eftekhar, Wenky Ma, Sukhvinder Kalsi-Ryan, José Zariffa

## Abstract

**Background:** Spinal cord injuries (SCI) can affect muscle control, often resulting in significant functional impairments. Damage to sensorimotor pathways in the spinal cord can lead to muscle weakness, which may gradually recover during the first year post-injury. Functional electrical stimulation therapy (FEST) aims to enhance muscle strength, particularly when natural recovery diminishes. However, the recovery profiles of individual muscles treated with FEST during the chronic phase are still not well understood. The primary objective of this study was to characterize the timing and magnitude of gains in individual muscle strength during FEST. The secondary objective was to identify factors predictive of the response.

**Methods:** In this cohort study, we examined the strength recovery profiles of 136 muscles treated with FEST, from 17 participants with cervical SCI at the chronic phase of the injury. Electrophysiology was conducted at baseline to assess corticospinal tract (CST) integrity and the excitability of the lower motor neuron pools using surface electromyography (sEMG).

**Results:** Our findings reveal that 69 muscles did not respond to the treatment, while 67 muscles showed a median strength increase of one muscle motor score (MMS). Among responder muscles, achieving a 1 MMS increase required approximately 59 days (19.2 FEST sessions). A prediction model highlighted key predictors of responsiveness to FEST, including baseline MMS, characteristics of lesion location/severity, and neurophysiological indicators of CST integrity.

**Conclusion:** Our findings underscore the need for refined guidelines in rehabilitation medicine, particularly for FEST in cervical SCI. The data demonstrates that achieving a 1-point increase in muscle strength requires ≈59 days and 19 FEST sessions. Further evidence also suggest that CST integrity and the responsiveness of lower motor neuron pools may influence FEST outcomes. These insights will enable more personalized and effective rehabilitation strategies, optimizing outcomes and resource allocation for individuals living with cervical SCI.

## 1. Introduction

Cervical spinal cord injuries (SCI) are common and may lead to impairments in upper limb function.^1^ At the chronic phase after the injury, regaining upper limb function is a priority for individuals with tetraplegia.^2,3^ Neuromodulation may help in increasing the sensorimotor recovery after SCI, especially when the natural recovery plateaus (> 6 months after the injury).^4^ For example, a recent safety and efficacy trial on non-invasive spinal cord neuromodulation indicated improvmeents in arm and hand function in chronic tetraplegia.^5^ Similarly, functional electrical stimulation has been used to improve the function of the upper extremities^6^ and functional electrical stimulation therapy (FEST) is among one of the most common neuromodulation techniques to assist in this regard.^7^

There is no consensus about the best duration and parameters of FEST, nor how long it takes to see a response in upper limb muscles, which can be characterized by an increase in muscle strength. In clinical trials aiming at understanding how FEST improves upper limb function post-SCI (*e.g*., reaching and grasping), FEST may employ sessions of 45-60 min (3-5 days/week), for 8-16 weeks, for a total of ≈ 40 sessions.^8^ Nonetheless, there is a lack of information about how FEST influences the motor recovery profile of individual muscles, how many days and session are needed to see an improvement in strength, and the magnitude of potential strength recovery. This is important because the recovery of motor strength is related to upper limb function since the pattern of residual strength in upper limb muscles can predict recovery of function after SCI.^9^ In actual clinical environments, FEST therapy schedules may be more variable than in research studies as a result of financial considerations, patient availability, and healthcare system capacity. In order to ensure that available therapy time is used effectively, it is important to understand the the magnitude and timing of the motor recovery with FEST.

Identifying factors that are predictive of therapeutic response is also necessary to support clinical planning. It has long been recognized that residual voluntary force production is indicative of favorable recovery prognosis during the natural recovery process after SCI.^10,11^ More recently, the pivotal role of corticospinal tract (CST) integrity in this process has also been emphasized.^12^ Recent evidence has also detailed the relationship between spasticity and CST integrity^13–15^,raising the possibility that the presence of spasticity may correlate with improved responsiveness to FEST.

Here, we present a detailed analysis of the recovery profiles of individual upper extremity muscles following FEST after cervical SCI. The primary objective of the study was to establish a dose-response relationship that could inform clinical guidelines. The secondary objective was to investigate baseline factors—both clinical and neurophysiological—that influence treatment response.

## 2. Materials and Methods

### Participants and assessments

Individuals with cervical injuries (C3-C6) in the chronic phase after the injury (> 6 months post-SCI) who were scheduled to receive FEST from a clinical program at our institution or through a clinical trial taking place at the time of this study were enrolled in our study (University Health Network REB approval numbers: 17-6029 and 19-5395.6).^16^ Additional participants were later recruited and received FEST through this study (Clinicaltrials.gov: NCT 05462925). The FEST instrumentation and protocols (described below) were the same in all cases. The research followed the Declaration of Helsinki and was approved by the Institutional Review Board of the abovementioned institution. Written consent was obtained from all 22 individuals enrolled in this study. Five participants did not complete the study, thus the final sample size used for this investigation was 17 individuals. We used the International Standards for the Neurological Classification of SCI (ISNCSCI) to evaluate the level and severity of injury.^17^ We assessed a total of 136 muscles from different upper extremity muscle groups including proximal upper extremity muscles (*Anterior* and *middle deltoids*, *Biceps brachii*, and *Triceps brachii*), forearm muscles (*Extensor carpi radialis* and *ulnaris*, *Extensor digitorum communis*, *Flexor digitorum superficialis*, and *flexor pollicis longus*) and intrinsic hand muscles (*Opponens pollicis*, *Flexor pollicis brevis*, *First dorsal interossei*, and *Abductor pollicis brevis*). Each participant underwent one baseline neurophysiological assessment before starting the course of therapy, with ongoing monitoring of MMS using MMT before each FEST session.

### Stimulation device and therapy

The delivery of FEST was accomplished using the MyndMove^®^ stimulation device (MyndTec, ON, Canada). Many stimulation protocols for the upper extremity were used, and included protocols specifically designed for individuals living with SCI. Specialized therapists who had undergone training specific to this device employed a variety of reaching and grasping movements, while using programmed stimulation sequences to facilitate functional movements of the arm(s) and/or hand(s). A detailed list of the protocols used in this study is available in Supplementary Table 1. The FEST was personalized to each participant based on the individual’s needs and goals. The FEST involved a different set of muscles for each protocol. The muscles stimulated during each protocol and the size of the stimulation electrodes were obtained from the manufacturer’s manual. Detailed information about the current delivered to each muscle and the time of treatment in each protocol was extracted from the reports generated after each FEST session. For each muscle in each session, the current delivered was multiplied by the number of cycles the protocol was administered. This resulted in the FEST dose delivered to each muscle *per* session in mA, which was summed across all FEST sessions for each muscle. Similarly, the time (in minutes) of FEST to each muscle was calculated.

### Manual muscle testing (MMT), muscle motor score (MMS)

Aligning with the clinical practice of FEST, the therapists selected muscles for assessment based on each patient’s specific needs and preferences to regain or improve necessary functions of the upper extremity., For the selected muscles, muscle strength was evaluated using Manual Muscle Testing (MMT).^17–19^ The MMT was scored using a standard 0-5 Medical Research Council (MRC) grading system^20^ by a trained physical or occupational therapist before each FEST session: 0 = Total paralysis; 1 = Palpable or visible contraction; 2 = Active movement, full range of motion with gravity eliminated; 3 = Active movement, full range of motion against gravity; 4 = Active movement, full range of motion against gravity and moderate resistance in a muscle-specific position; 5 = (Normal) active movement, full range of motion against gravity and full resistance in a functional muscle position expected from an otherwise unimpaired person.^21^ For assessing muscle strength (muscle motor score; MMS) of muscles not included in the ISNCSCI we used the protocols described in the Graded Redefined Assessment of Strength, Sensibility and Prehension (GRASSP).^22^ For muscles without a defined MMT protocol in the ISNCSCI nor the GRASSP, the therapist team developed custom positioning and stabilization protocols to consistently apply the MRC scale. The analysis reported herein focuses specifically on strength recovery trajectories of individual muscles (segmental strength recovery), with the intent of supporting therapy planning; global and functional FEST outcomes have been the focus of previous studies.^23^

The MMT was conducted before every FEST session by the treating therapist. On days when an assessment was missed, for example due to time constraints (36/512 = 7% of total FEST sessions), the MMT values were carried forward from the most recent available clinical measurement to ensure a continuous recovery profile. For analyses comparing recovery profiles of muscles from individuals with different durations of therapy, the final available value was also carried forward. Typically, eight to ten muscles were evaluated per participant.

### Neurophysiology – volitional neuromuscular activation

In order to gain insight into the influence of CST integrity on FEST recovery profiles, we analyzed surface electromyography (sEMG) during voluntary contractions. This choice of modality was motivated by the prospect of integrating our findings into technology available to therapists at the point of care in the future, which would be more difficult if relying on transcranial magnetic stimulation to elicit motor evoked potentials. The sEMG was conducted before the start of the FEST (baseline). sEMG signal from all upper limb muscles tracked in this study was acquired by an 8-channel Bagnoli^®^ system (Delsys, MA, USA) with a 4kHz sampling frequency and 1000X amplification (hardware filter at 20-450Hz), using LabChart (AD instruments, Colorado Springs, USA). The bipolar differential dry electrodes (Delsys, DE 2.1 EMG Sensor) were positioned over the muscle belly using double-sided adhesive tape after removing any excess hair and cleansing with alcohol. The positioning and orientation of the electrodes were based on the SENIAM guidelines^24^ unless guidelines were not available for a specific muscle group.^25^ sEMG signals were acquired during isometric voluntary attempts using the MMT assessment protocol for each muscle (see above). The resting-state sEMG was acquired for 30 seconds with the participant relaxed before the muscle contractions. The resting-state sEMG protocol was followed by three trials of 5-second maximum voluntary contractions with a minimum of 30 seconds between contractions. All sEMG metrics were averaged over 3 trials of the maximum voluntary contraction. sEMG was filtered using a 2^nd^ order Butterworth filter, band-pass between 20-450 Hz, and a band-stop filter between 59-61 Hz (to remove the power line interference). The amplitude of the sEMG was quantified using the root mean square (RMS) of the filtered sEMG signal for the entire analysis window during maximal voluntary contractions. The analysis window was determined through visual inspection and included the period from the onset to the offset of voluntary modulation of the sEMG interference signal. This process was also guided by an external trigger signal (square waveform), which was recorded by the AD converter and timed by the neurophysiologist during acquisition. The timing was based on verbal commands indicating the initiation of maximum voluntary contraction and the subsequent command to cease the contraction.

The median frequency was calculated using a procedure previously described.^26^ The spectral profile of each window was divided into 100 bins (2.5 Hz resolution) and the numerical integral was calculated. The integrated sEMG was then split into two halves of equal areas, this value (split point) constituting the median frequency (MDF) in Hz. To visualize the frequency component of the sEMG throughout the volitional contraction, we computed the short-time Fourier transform-based spectrogram of sEMG signal (automatic window length with 75% of overlap).

### Neurophysiology – spontaneous MU firing at rest

The sEMG dataset described above was further analyzed to characterizing spontaneous MU firing at rest. All post-processing of the sEMG data was performed using cutom build software and graphical user interfaces using LabVIEW Version 20.0.1 (National instruments, Texas, USA). The sEMG signals during resting intervals were visually inspected to identify spontaneous MU firing. This assessment included periods when the participant was relaxed before muscle contractions and during 30-second rest intervals between contractions. In some instances, unit spasms occurred shortly after voluntary contractions. In other words spontaneous MU firing at rest was quantified at: (1) the resting trial prior to maximal voluntary contractions, (2) or the relaxation period preceding the maximum voluntary contraction for the target muscle group, and (3) or the resting intervals between the maximum voluntary contraction. Segments of the sEMG signal containing spontaneous MU firing were isolated and cropped based on the duration of the unit spasms. A visual threshold was applied to differentiate MU spikes from background sEMG noise using a custom graphical user interface. A peak detection function, using the established visual threshold, was employed to segment each MU spike for further analysis. The threshold was set visually based on the sEMG interference signal to identify both large and smaller amplitude spikes while avoiding noise interference. The assessor monitored the number of detected peaks in real time and made informed decisions based on this ongoing assessment. From each individual waveform, we extracted 13 features: kurtosis, skewness, range, RMS, variance, index of maximum, index of minimum, maximum value, minimum value, mean, standard deviation (SD), median, and mode. Principal component analysis (PCA) was used to plot each MU spike in a latent space, retaining 2 principal components (fixed parameter in the model). Subsequently, k-means clustering was performed with random initialization, utilizing 3 clusters, 50 iterations, and exhaustive hyperparameter search, with evaluation based on the Jaccard index. The resulting cluster labels were used to categorize the 3 classes of MUs across the continuous sEMG record. The number of clusters was a fixed parameter in the clustering model, determined based on prior research that identifies a limited number of spontaneously firing motor units in individuals with post-SCI conditions.^27–31^ Quantitative measures included MU amplitude (peak-to-peak, in mV), firing frequency (in Hz), interspike interval (in ms), and the coefficient of variation (CoV) of the interspike interval of each MU.

### Recovery profiles

To understand the change in MMS after treatment, MMS change (ΔMMS) was calculated by subtracting baseline strength (the average of the first two MMS scores) from endpoint strength (the average of the last two MMS scores). Considering that most clinical studies show long-term effects of electrical stimulation (13 days to 6 weeks) on muscle strength^32^, this averaging procedure was conducted to account for variability in the initial and final assessments. The motor recovery (%) is the sum of the motor score of all upper limb muscles included in the assessment *per* participant, normalized to the maximum this score could reach (ceiling: considering each muscle can achieve a maximum of 5 points). Then this score was expressed as a percentage change from baseline at the participant level.

We categorized muscles as therapy responders or non-responders depending on whether a longitudinal increase in strength was observed over the course of the FEST sessions. Considering the complex and variable responses of muscles to FEST, we utilized the expertise of four therapists involved in the study to determine each muscle’s response to therapy (SK, WM, AC, PE). Using a majority vote approach, therapists were presented with comprehensive recovery profiles for all 136 muscles, presented graphically. To maintain objectivity, therapists were blinded both to patient identities and to each other’s assessments. Each therapist independently evaluated whether a muscle qualified as a responder or non-responder and indicated the timing of recovery onset. We used the majority vote outcomes to stratify into responder and non-responder groups—ties (2 instances) were broken by GB.

### Statistical analysis

Statistical analyses were performed using Statistical Package for the Social Sciences (SPSS) v.25 and GraphPad Prism v.9. The unit of analysis was the individual (17 participants), the muscle (136 muscles) or the MU (216 MUs). Descriptive statistics were presented as mean ± SD or median with interquartile range. Normality of data was assessed using the Shapiro-Wilk test, and parametric or non-parametric tests were applied accordingly. To determine the timing of recovery with FEST between responder and non-responder muscles, multiple Kolmogorov-Smirnov tests with the Holm-Šídák method (α = 0.001) were conducted to adjust for multiple comparisons. For assessing factors contributing to individual muscle response to FEST while accounting for muscle dependency (multiple muscles from the same participants), generalized estimating equation models (ordinal logistic) were employed. Participant ID was included as a subject variable, and muscle ID as a within-subject variable. Predictors for FEST response included American Spinal Injury Association (ASIA) Impairment Scale (AIS), the distance from the neurological level of injury to the muscle myotome (DST), myotome, baseline MMS, baseline volitional sEMG amplitude (root mean square), and baseline volitional sEMG frequency (median frequency), with age as a fixed covariate in the model. Continuous neurophysiological data were transformed into binned ordinal data: sEMG amplitude into a scale of 0-18 and sEMG frequency into a scale of 0-49. Model effects were visualized for the aforementioned factors using type I statistics and the Wald chi-square method. For the correlational analysis, we used Spearman correlations and the unit of measure was the individual muscle. Significance was set at α = 0.05 in all instances if not specified above.

## 3. Results

A hundred and thirty six muscles from 17 participants were assessed in this study (**Table 1**). The number of FEST sessions (11-40) and duration of treatment (41-246 days) varied among participants. The sample was predominantly composed of males and the majority (10/17) were motor incomplete participants. There was a mean ± SD of 8 ± 0.8 upper limb muscles assessed per participant.

**Table 1.**
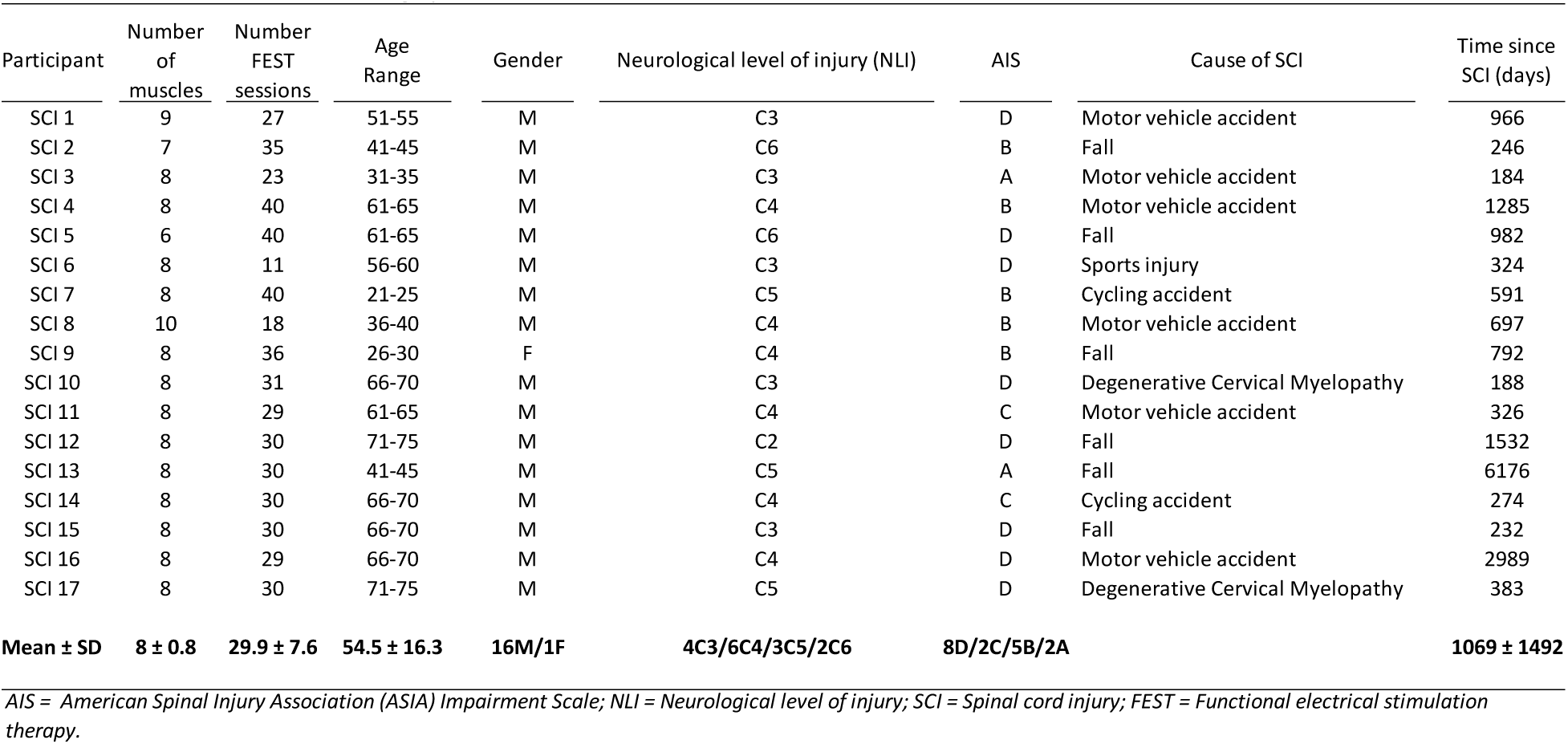
Patient Characteristics and Demographic Information.

### Characterization of recovery profiles

The recovery profile of each participant is described in **Figure 1**, noting the variability in the duration of treatment and variable response in the motor recovery. Some participants presented a fast response to the treatment, and others a more gradual change.

**Figure 1.**
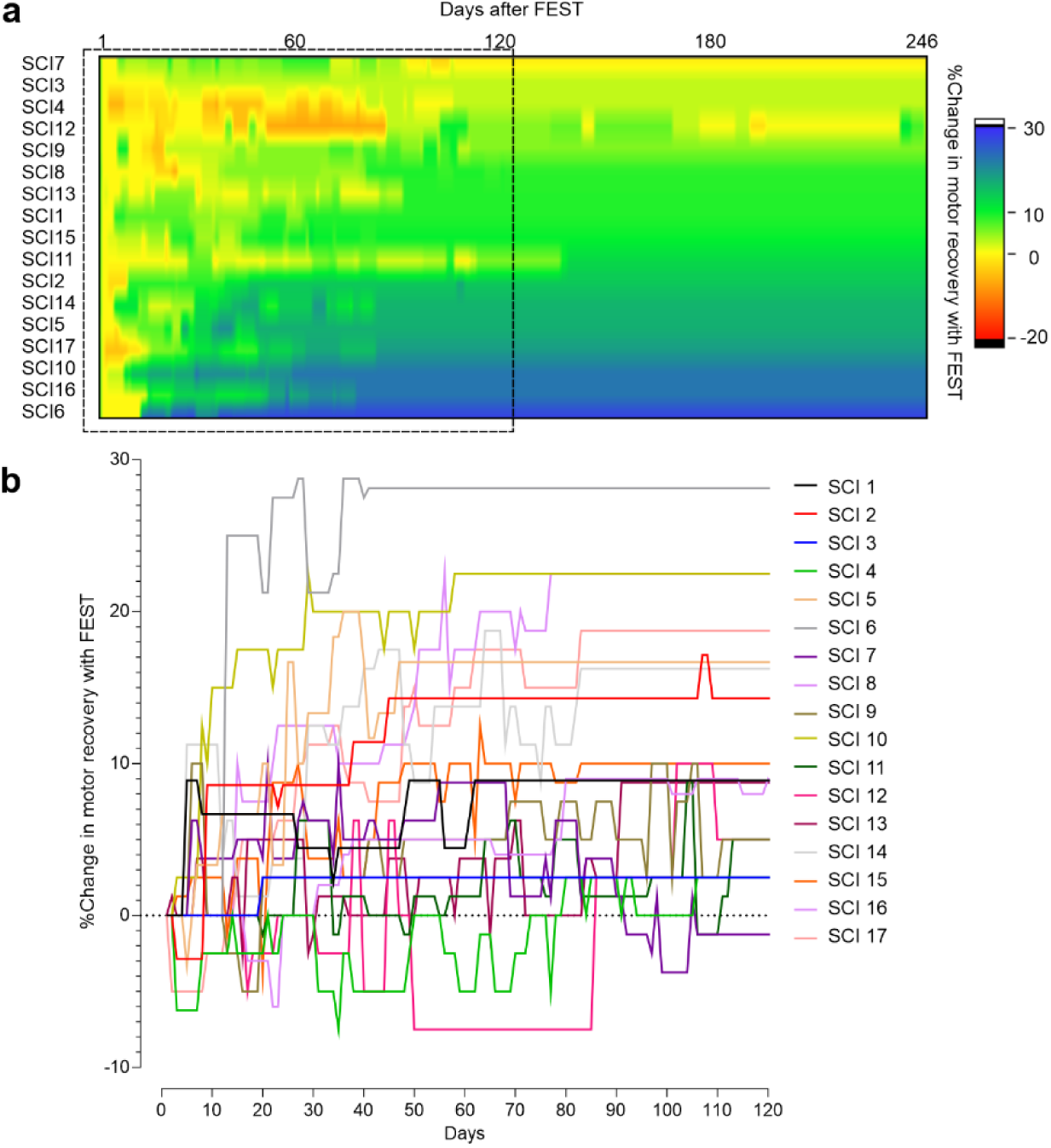
Motor Recovery Profiles with Functional Electrical Stimulation Therapy (FEST) in Chronic SCI. (**a**) Heat map depicting the cumulative Muscle Motor Scores (MMS) across all tracked muscles for each participant, normalized to baseline (% Change in motor recovery with FEST) throughout therapy. (**b**) Temporal progression of the data shown in (**A**), plotted from day 0 to 120, highlighting the therapeutic window where significant gains with FEST were observed. *FEST = Functional Electrical Stimulation Therapy; SCI = Spinal Cord Injury*.

The recovery profiles of individual muscles are depicted in **Figure 2**. Variability in strength response was observed throughout the course of FEST, both over time and between muscles. Among the 136 muscles analyzed, 67 showed strength improvement during the FEST period (majority vote procedure).

**Figure 2.**
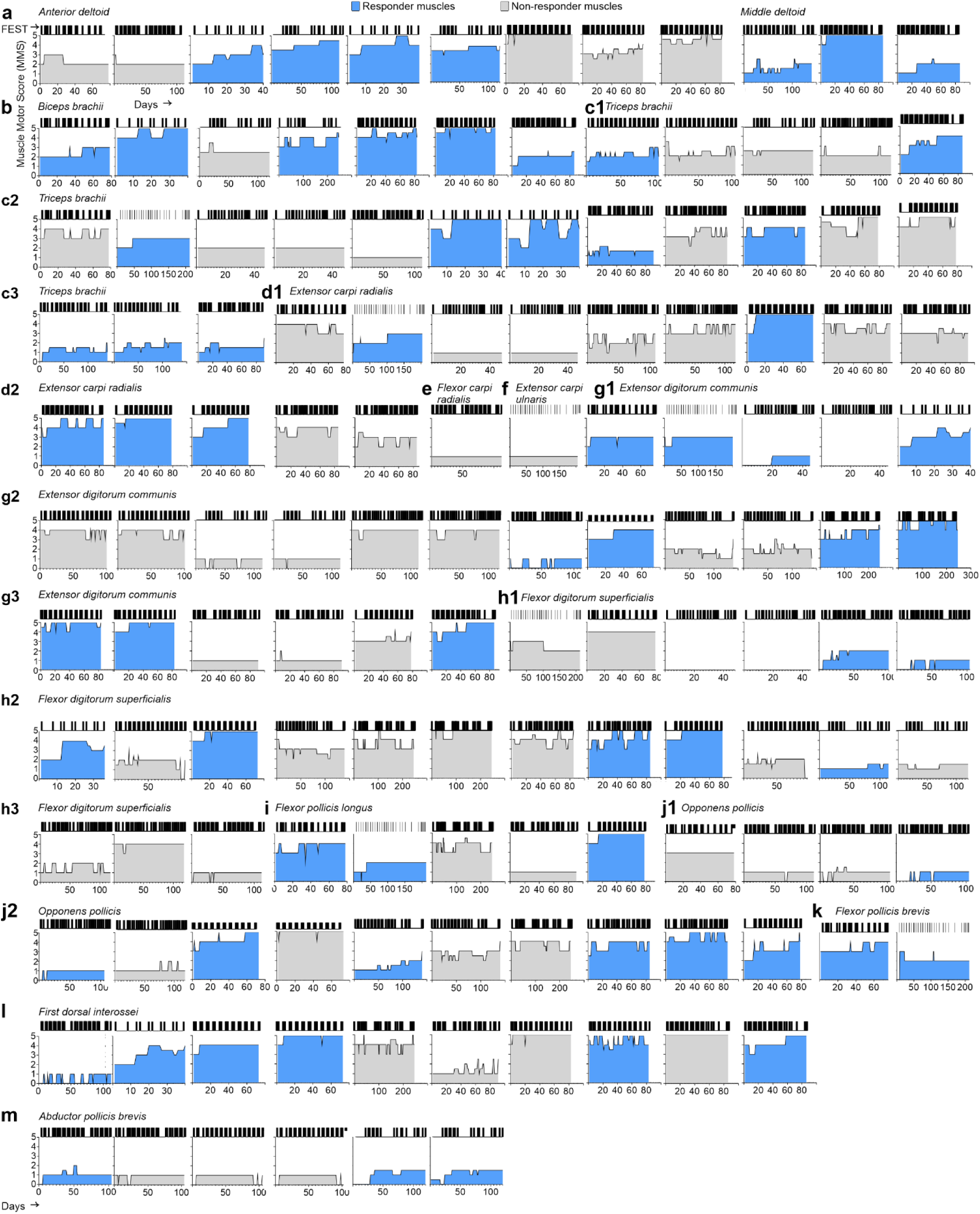
Muscle Motor Score (MMS) Recovery Profiles of the 136 muscles monitored throughout the Functional Electrical Stimulation Therapy (FEST) treatment. (**a**) *Anterior* and *middle deltoid*. (**b**) *Biceps brachii*. (**c**) *Triceps brachii*. (**d**) *Extensor carpi radialis*. (**e**) *Flexor carpi radialis*. (**f**) *Extensor carpi ulnaris*. (**g**) *Extensor digitorum communis*. (**h**) *Flexor digitorum superficialis*. (**i**) *Flexor pollicis longus*. (**j**) *Opponens pollicis*. (**k**) *Flexor pollicis brevis*. (**l**) *First dorsal interossei*. (**m**) *Abductor pollicis brevis*. Each graph illustrates the muscle motor score (MMS) recovery profile with Functional Electrical Stimulation Therapy (FEST). *The hash marks above each graph indicate days when FEST was administered. Graphs marked in blue indicate muscles classified as responders by therapist assessment. MMS = Muscle Motor Score; FEST = Functional Electrical Stimulation Therapy*.

The median muscle recovery profile of responders and non-responder muscles is shown in **Figure 3a**. Note the fast response of most responder muscles compared to the non-responder muscles. It took a median of 25 days and 7.64 FEST sessions for responder muscles to achieve an 0.5-point increase in MMS (Kolmogorov-Smirnov test to compare cumulative distributions; p < 0.001; **Figure 3b-3c**). A 1 point in MMS gain was evident after 59 days/19.2 FEST sessions. When considering all muscles (responders and non-responders), there was a 0.5-point significant increase in strength with FEST [Wilcoxon test, W = 4544, p < 0.0001; number of pairs = 136, number of ties (ignored) = 32; **Figure 3d-3e**]. **Figure 3f-3g** show a heat map of the recovery profile (ΔMMS) with FEST for responder and non-responder muscles during the 120 initial days of FEST.

**Figure 3.**
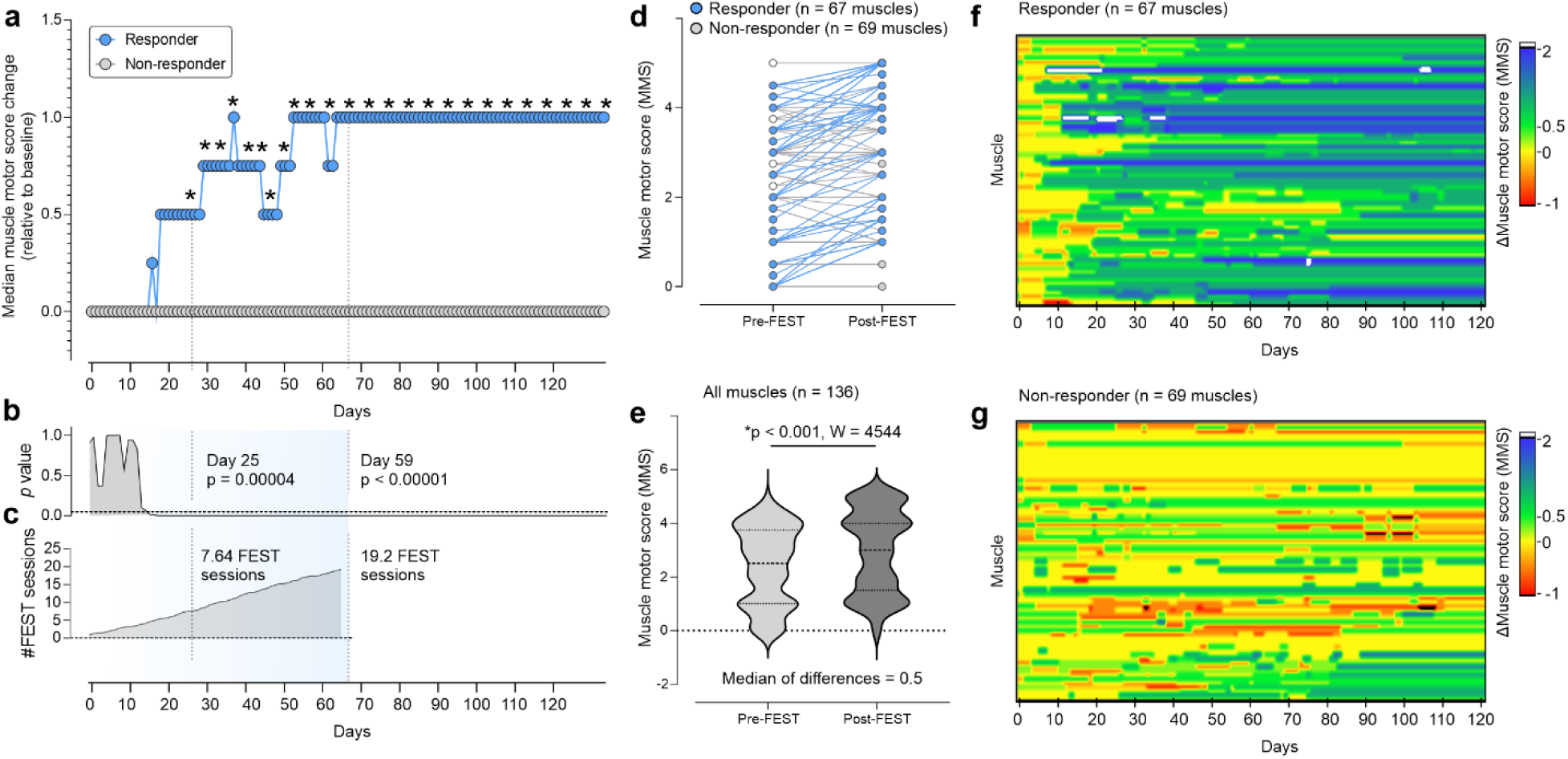
Recovery profiles of individual upper limb muscles with FEST. (**a**) Median muscle motor score (MMS) change (relative to baseline MMS); (**b**) *p* values form the Kolmogorov-Smirnov test used to compare cumulative distributions of responder and non-responder muscles throughtout the course of the FEST; and, (**c**) the cumulative number of FEST sessions to observe the effect on MMS. (**d**) Change in MMS across all 136 muscles between pre-FEST (baseline) and post-FEST (endpoint). (**e**) A significant 0.5 MMS increase was evident when comparing baseline to endpoint (Wilcoxon test, p < 0.001). (**f**) Heat map showing the ΔMMS thought out the FEST course for responder and (**G**) non-responder muscles. *Data is median or median ± 95% interquartile interval. MMS = muscle motor score; FEST = Functional electrical stimulation therapy*.

### Factors predictive of response

To explore what baseline factors could predict responsiveness to FEST, our initial focus was on CST integrity, as reflected by sEMG during the maximum voluntary contractions of the target muscles (see **Figure 4a**). A correlational analysis indicated that the severity of the lesion (AIS grade) was positively correlated with ΔMMS with FEST (Spearman correlation, p < 0.001, r = 0.399). Individuals with less severe lesions also exhibited greater residual strength (MMS; Spearman correlation, p < 0.001, r = 0.511) and volitional drive (sEMG amplitude; Spearman correlation, p = 0.001, r = 0.287). This suggests greater CST integrity for muscles in individuals classified as AIS C and D (**Figure 4b**).

**Figure 4.**
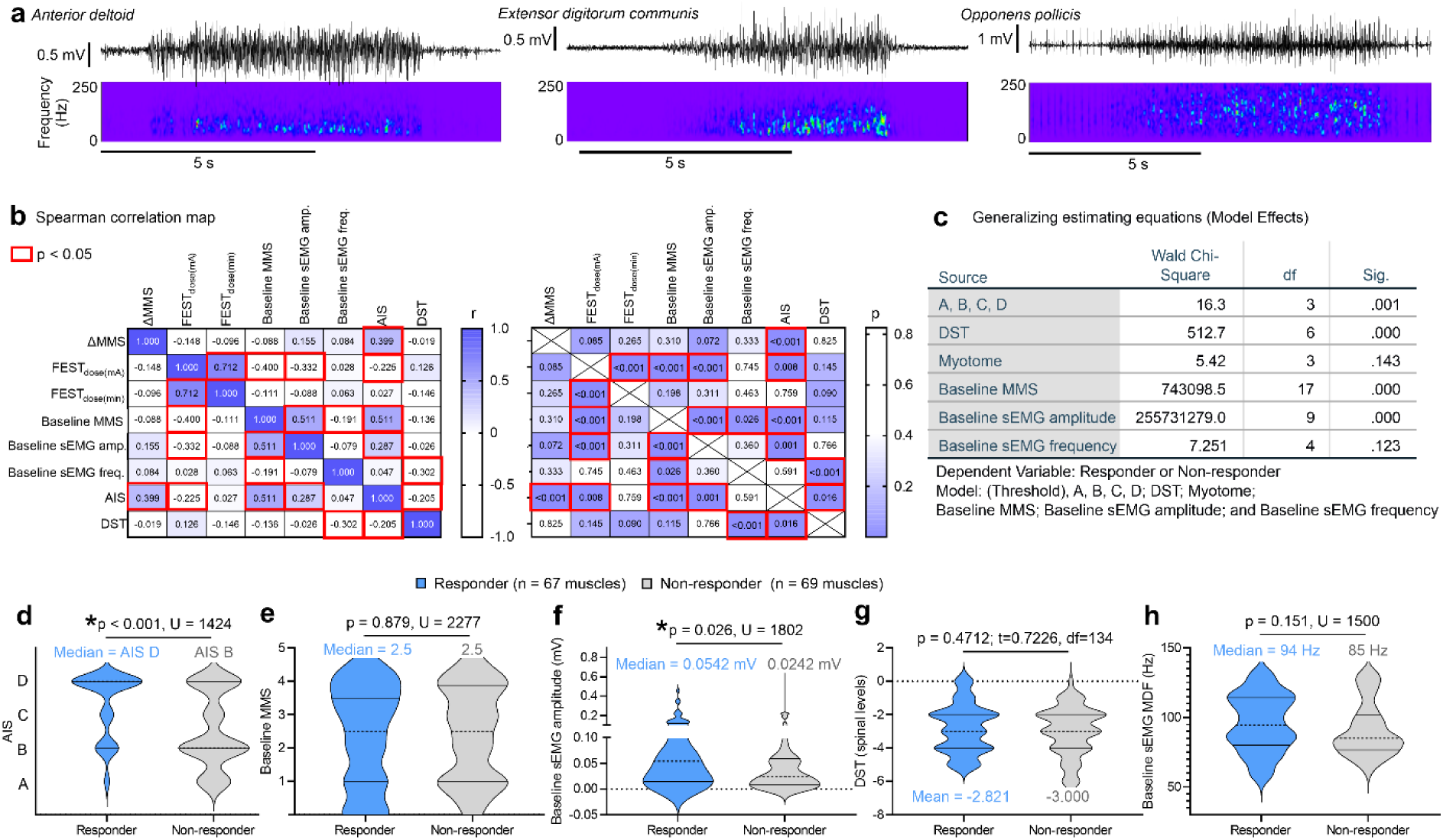
CST integrity is pivotal for the recovery of individual upper limb muscles with FEST. (**a**) Examples depicting neurophysiological testing using surface electromyography (sEMG) to assess corticospinal tract (CST) integrity. Participants performed maximum voluntary contractions to measure lower motor neuron activation by descending projections. (**b**) Correlational analysis showing the relationship between lesion severity/completeness and changes in Manual Muscle Score (ΔMMS) with Functional Electrical Stimulation Therapy (FEST). Greater lesion severity correlated positively with ΔMMS. Participants with less severe lesions demonstrated higher residual strength (Baseline MMS) and volitional drive (Baseline sEMG amplitude), suggesting greater CST integrity. (**c**) Generalized estimating equations models investigating baseline factors predicting muscle responsiveness to FEST. Results highlight the significance of AIS classification, baseline strength (baseline MMS), and baseline sEMG amplitude in predicting treatment response. (**d**) Distribution of responder and non-responder muscles based on AIS classification (Mann Whitney test, p < 0.001). Responder muscles predominantly from AIS D individuals. (**e**) Similar baseline MMS levels observed around 2.5 for both groups. (**f**) Muscles with greater CST integrity (Baseline sEMG amplitude) show a more recovery (Mann Whitney test, p = 0.028). (**g**) No apparent differences in the recovery profile pattern with the distance from level of injury (DST). (**h**) Substantially greater recovery for muscles with high frequency components is evident when plotting the recovery profile in relation to baseline sEMG frequency. Muscles with residual activation of faster, higher threshold motor units (MUs) seem to respond better to FEST *Data is mean or median ± 95% interquartile interval. MMS = muscle motor score; FEST = Functional electrical stimulation therapy; MU = motor unit; CST = corticospinal tract; DST =-distance from the level of injury; AIS = American spinal injury association (ASIA) impairment scale*.

The correlational analysis indicated a few other associations important to note. In line with previous findings^33^, there was a correlation between baseline MMS and sEMG amplitude (Spearman correlation; p = < 0.001, r = 0.511). The dose of FEST (in mA) delivered to each muscle was correlated with the baseline MMS (Spearman correlation; p < 0.001, r = -0.400), sEMG amplitude (Spearman correlation; p < 0.001, r = -0.332), and lesion severity (AIS; Spearman correlation; p = 0.008, r = -0.225). These negative correlations indicate the delivery of most of the FEST dose to individuals and muscles with pronounced weakness at baseline.

We then employed generalized estimating equations models to investigate which baseline factors predict muscle responsiveness to FEST, accounting for dependencies within the dataset (since muscles are from the same individual). This analysis highlighted the significance of AIS classification, baseline strength (baseline MMS), and baseline sEMG amplitude in predicting treatment response (**Figure 4c**). Muscles from individuals with less severe SCIs closer to the lesion level (DST), with greater residual strength (Baseline MMS), and higher sEMG amplitudes demonstrated better responses to FEST (responder muscles). Further descriptive analysis at the muscle level revealed that responder muscles predominantly belonged to individuals classified as AIS D (non-responder muscles belonged to AIS B; Mann Whitney test, p < 0.001, U = 1424; **Figure 4d**), exhibited similar baseline MMS levels around 2.5 (**Figure 4e**), showed enhanced CST integrity (sEMG amplitude; Mann Whitney test, p = 0.028, U = 1802; **Figure 4f**), and were located at a comparable distance below the injury (DST; **Figure 4g**). Additionally, responder muscles exhibited evident and substantial activation of faster, higher threshold motor units, as indicated by substantially higher sEMG frequencies at baseline compared to non-responder muscles (Mann Whitney test, p = 0.151, U = 1500; **Figure 4h**). This underscores the importance of CST integrity in facilitating muscle response to FEST.

### Spasticity analysis

To investigate the interplay between spasticity, CST integrity, and motor recovery with FEST, we employed neurophysiology to measure the spontaneous firing of MUs (unit spasms) at rest, serving as an indicator of involuntary muscle activation, similar to that observed during spastic events (examples of spontaneous MU firing at rest are shown in **Figure 5a**). Spontaneous firing of motor units (MUs) was observed in 34 responder and 38 non-responder muscles. For muscles exhibiting MU firing at rest, we implemented an MU extraction procedure using amplitude thresholding to detect these events (see example in **Figure 5b**), followed by dimensionality reduction and clustering (see **Figure 5c-g**). We identified 102 motor units from responder muscles and 114 from non-responder muscles. Responder muscles tended to show higher MU amplitudes during these events compared to non-responder muscles (Mann Whitney test, p = 0.243, U = 5279; **Figure 5h**). However, no significant differences or apparent trends were found in terms of the frequency of MU firing, interspike interval, or its variability (**Figure 5i-k**).

**Figure 5.**
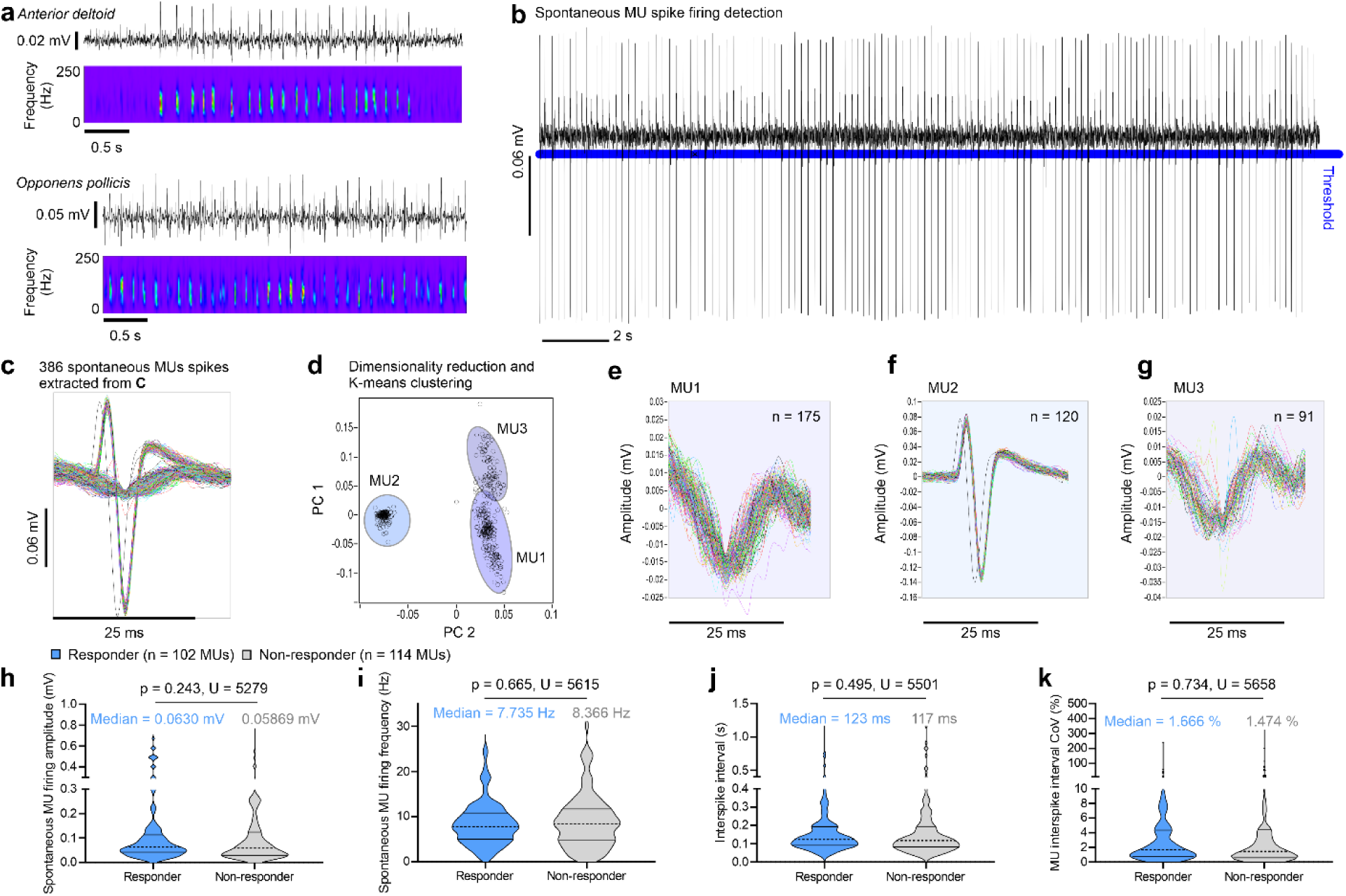
Spasticity and Motor Recovery in Functional Electrical Stimulation Therapy (FEST). (**a**) Illustration depicting examples of spontaneous firing of motor units (MUs) at rest, observed as unit spasms, which indicate involuntary muscle activation akin to spastic events. (**b**) Demonstration of the MU extraction procedure using amplitude thresholding to detect spontaneous MU firing events in muscles. (c,d) Dimensionality reduction and clustering of MU firing patterns (examples of extracted MU waveforms shown in panels **e-g**). (**h**) Comparison of MU amplitudes during spontaneous firing events between responder and non-responder muscles. Responder muscles exhibited substantially higher MU amplitudes, although the difference was not statistically significant (Mann Whitney test, p = 0.243, U = 5279). (**i-k**) Analysis of MU firing characteristics including frequency, interspike interval, and variability, showing no significant differences or discernible trends between responder and non-responder muscles. *Data is mean or median ± 95% interquartile interval. FEST = Functional electrical stimulation therapy; MU = motor unit*.

## 4. Discussion

In this study, we provided a comprehensive analysis of individual upper extremity muscle recovery patterns following FEST, and investigated key baseline factors that influence treatment response. Data collection followed a pragmatic approach that reflected FEST protocols in use at our institution, in order to support the clinical applicability of the findings. Our results highlight that muscles affected by less severe initial lesions displayed more pronounced improvements with FEST. Among muscles categorized as responders, achieving a gain of 1 point on a 6-point Likert scale (MMS) took about 59 days or 19.2 FEST sessions. Additionally, we found that muscles retaining some integrity of CST projections, assessed *via* volitional sEMG), exhibited heightened responsiveness to the therapy. Moreover, preliminary findings indicate that individuals presenting clinical signs of upper extremity spasticity or spontaneous motor unit (MU) spasm at rest benefited substantially from FEST. These findings establish a compelling connection between CST integrity and therapeutic outcomes in FEST, suggesting potential avenues for tailored treatment approaches guided by neurophysiological markers.

This study represents the first detailed examination of motor recovery at the muscle level using FEST in individuals with cervical SCI. Previous research has described the effects of FEST on upper limb function, showing notable improvements such as enhanced voluntary grasping beyond conventional therapy effects.^34^ Similarly, series of 40 sessions of FEST demonstrates comparable effectiveness to traditional therapy in generating significant functional enhancements that endure beyond the treatment period.^23^ Given that upper limb function can be predicted by the combined strength of upper limb muscles in acute SCI (1-6 months)^9^, it was anticipated that the enhancement of function with FEST at chronic stages would be supported by the strength recovery of individual upper limb muscles. It is important to note the difference of previous studies, which, conversely to the present study conducted the analysis in the acute phase of SCI (< 6 months post-SCI)^9,34^ or had unbalanced FEST and conventional therapy groups.^23^ This may have overlapped with spontaneous recovery periods that are characterized by pronounced gains in sensorimotor function, especially in less severe cases of SCI. Therefore, the gains with FEST presented here may be understood under the perspective of the natural recovery plateau occurring 6 months after the injury.^10,35–41^ In the chronic phase of SCI, even small gains are capable to promote enhanced function and quality of life after SCI, particularly in regaining hand function—a critical priority in cervical SCI rehabilitation.^42^ Our results are also important under the perspective of extensive effort being devoted to the understanding recovery of upper extremity function^10,11,36–38,43–47^. However, less attention has been given to the recovery of individual muscles (segmental recovery) through neurorehabilitation following SCI.^12,35^ Segmental recovery and its prediction are a current focus in the field of SCI. ^12,48,49^

Expert therapists rely on several key considerations when evaluating muscle motor recovery following FEST. Ensuring consistency in MMT scores provides a reliable measure of motor improvement. Support systems and resource availability outside therapy sessions play a significant role, potentially influencing recovery positively or negatively. Variability in FEST duration and frequency among participants, along with the timing of assessments and sessions throughout the day, further shape outcomes. Considerations for scale non-uniformity and qualitative feedback are vital for comprehensive evaluation and treatment optimization. A detailed summary of the therapists’ perspectives on assessing muscle responsiveness to FEST is presented in the Supplementary Material.

The main factors indicating the recovery of muscle strength with FEST are long known predictors of natural recovery after the lesion. For example, the residual baseline strength indicates the amount of residual sensorimotor control over muscles affected by the lesion and is a positive predictive factor after SCI.^50^ Along these lines, the AIS classification – which is also quantified using the above-mentioned residual strength to muscles, is a powerful predictor of post-SCI recovery.^43,50^ All of these predictors of FEST responsiveness are somewhat related to the integrity of CST projections, which is the main source of control for upper extremity muscles—especially those controlling dexterous hand movements.^12,51,52^ Neurorepair of CST fibers is most likely to occur close to the lesion location, as these newly formed axons have limited neuroregeneration capacity.^53^ Muscles that are 1 to 3 levels below the lesion are more likely to have some degree of partial innervation (zone of partial innervation) and show greater recovery after SCI.^43,54^ On the other hand, segments closer to the lesion are more likely to be associated with lower motor neuron injuries^55^, such that the relationship between DST and responsiveness to FEST may be subject to competing factors. Overall, our results support the responsiveness to FEST in muscles from individuals with less severe lesions (AIS classification), with greater CST integrity, and with a closer distance from the level of injury.

Baseline neurophysiological biomarkers may also add to the prediction of responsiveness to FEST. In this regard, the most prominent result of the present study indicates that the volitional drive captured by sEMG during maximum voluntary contractions has a strong value in predicting muscle recovery after FEST. Additionally, preliminary results suggest that unit spasms present at rest^27^ may influence the response to FEST. These spasms do not appear to hinder the effectiveness of the therapy, conversely, trends indicate that they may be associated with a better response. Further research is needed to determine if spasticity could serve as a biomarker for predicting response to FEST. Some motor units may show prolonged, at rest, or contraction-induced firing after the voluntary contraction^27–31^. This spontaneous or induced MU firing (unit spasms) may last for minutes.^28,29^ It has been suggested that the active source of such unit spasms is the motoneurons themselves^29,30^ and that additional inputs to lower motor neuron may facilitate functional firing of residual MUs, likely reflecting neuroplasticity within the MU circuit with sensorimotor recovery from SCI.^56^ We investigated if weakened or paralyzed muscles displaying MUs that fire spontaneously would have a possible influence on MU contractile properties after SCI^28,29^—influencing the response to FEST. MU firing characteristics in individuals with chronic cervical SCI was previously studied, with reports of MUs that continued firing involuntarily after cessation of voluntary contraction (at frequencies around of 5.9 Hz – 15.4 Hz).^29,31^ The underlying mechanisms for this activity remain unclear. In a more in-depth study of the properties of these spontaneously firing MUs in SCI, it has been suggested that regularly firing MUs are more excitable—and produce more force^31^, compared to irregularly firing units. Here we show that spontaneous MU firing at rest occurred at a similar frequency range (7.7 Hz and 8.4 Hz for responder and non-responder muscles, respectively). We did not observe any differences in terms of regularity of firing in MUs from responder and non-responder muscles, nonetheless, there was a substantial increased amplitude in responder muscles. The spontaneous regular firing of motor units may indicate active intrinsic properties, such as persistent currents within motoneurons, which are evident even in the absence of voluntary drive.^31^ Considering this potential, it is plausible that muscles demonstrating stronger spontaneous MU activity may be more responsive to additional sensorimotor feedback, such as that provided by FEST, which could enhance their functional recovery.^56^ Further studies are necessary to investigate the relationship between spasticity^13–15^, unit spasms^29,31^ and neurorecovery after FEST. Finally, future studies should approach the use of biomarkers from sEMG to predict the responsiveness to FEST, including detailed MU decomposition using high density sEMG, time- and frequency-domain analysis and additional electrophysiological techniques (such as motor-evoked potentials).^12,57–60^

### Limitations (107 words)

This study has a few limitations: (1) Given the pragmatic nature of the study design, our participant sample was heterogeneous. This variability, combined with the relatively small sample size, limits a more definitive analysis of the factors contributing to FEST response. On the other hand, pragmatic study designs can provide findings that generalize better to real-world clinical settings than those of clinical trials with strict inclusion criteria^61^. Furthermore, the focus on individuals in the chronic stage after injury helps to ensure that observed changed were causally related to the FEST. (2) The gender imbalance, which reflects the high male-to-female ratio (5:1) commonly observed in traumatic SCI.^62^ (3) The reliance on manual measures of muscle strength provides relatively coarse information in comparison to sensor-based methods. Nonetheless, MMT is a pragmatic approach that provided the flexibility to assess different muscles for each person depending on their treatment goals, and could be performed quickly without interfering with the therapy sessions. (4) The FEST dose recorded may only partially represent the actual therapy delivered, as data logs did not capture the full extent of the FEST sessions. (5) Spasticity assessments were a secondary analysis based on available electrophysiolgical data without following specific protocols like the Modified Ashworth scale, which are not designed to assess any individual muscle.^63^ Future research should employ more rigorous protocols, including neurophysiological assessments using sEMG to quantify properties reflecting lower motor neuron spontaneous activity (reviewed in^59^).

## 5. Conclusion

In individuals with chronic SCI, FEST applied to the upper limbs typically results in modest and variable improvements in muscle strength. Sixty-seven of 136 (49.3%) muscles in our sample responded to FEST, with 19.2 FEST sessions being required to detect a 1-point difference in the MMS gain between responder and non-responder muscles. Key determinants of therapy response include injury severity and the integrity of the CST. Preliminary findings suggest that neurophysiological biomarkers, such as the magnitude of the volitional drive—a surrogate for CST integrity— has a pivotal role in predicating responsiveness to FEST. Assessing CST integrity at the muscle level could therefore aid in predicting which muscles are likely to benefit from FEST, facilitating targeted neurorehabilitation and optimizing therapeutic outcomes. These findings underscore the need for updated guidelines in rehabilitation medicine regarding the use of FEST for cervical SCI.

### Transparency, Rigor, and Reproducibility

The results reported in this article include participants who received FEST from 3 sources, all using the same instrumentation and training protocols. (1) A clinical program at our institution. In this case, the therapy was delivered clinically and the study team conducted additional sEMG and strength testing assessments. (2) A FEST clinical trial taking place at our institution at the time of this study (Clinicaltrials.gov: NCT 03439319). The team for the present study conducted additional sEMG and strength testing assessments. (3) Later, to increase the sample size and as a result of available funding, additional participants were recruited whose therapy was delivered by the same clinical team but funded through this study (Clinicaltrials.gov: NCT 05462925). Regardless of the therapy delivery scenario, all of the data reported in this article were collected under protocol 19-5395 approved by the Research Ethics Board of the University Health Network. All participants provided written informed consent. The device used to deliver the intervention is available commercially (MyndTec Inc., Toronto, Canada). The total target sample size was 30 participants x 6 muscles each, for a total of 180 muscles. Partly as a result of the pandemic, only 22 participants were enrolled. 5 did not complete the study, leading to a final sample size of 17 participants. However, we were able to track more muscles per participant than originally projected, partially compensating for the lower number of individuals and resulting in 136 muscles for analysis. The analysis plan for this study was not formally pre-registered. The source data for every figure is provided in the supplementary material for reproducibility of the analyses. Additional data can be made available upon reasonable request and completion of a data sharing agreement.

## Data Availability

Data can be made available upon reasonable request and completion of a data sharing agreement.

## Acknowledgments

This work was supported by the Wings for Life Spinal Cord Research Foundation (Project #210). We would like to thank MyndTec Inc. and their sponsored clinical trial “Restoration of Reaching and Grasping Function in Individuals With Spinal Cord Injury Using MyndMove^®^ Neuromodulation Therapy” (USAMRAA CDMRP-SCRIP – Protocol SC150251, Clinicaltrials.gov: NCT 03439319) for access to individuals receiving FEST. We would like to thank Dr. Mohammad Alavinia for his statistical assistance, as well as the individuals receiving FES therapy. We extend our gratitude to Dr. Cindy Gauthier and Cynthia Ho for their invaluable assistance in delivering specialized therapy.

## Author contributions

All authors contributed to writing and revising the manuscript. Therapy was administered by SK, PE, WM, and AC, with supervision by SK. Signal acquisition and analysis were conducted by GL and GB. Conception, funding, and overall supervision were provided by JZ.

## Supplementary Material

### Comprehensive Perspectives on Evaluating Motor Recovery After FEST

When evaluating the response of muscles to Functional Electrical Stimulation Therapy (FEST), several factors guide therapist assessment. Consistency in Manual Muscle Testing (MMT) scoring is crucial, providing a reliable measure of motor recovery. However, several factors may lead to the lack of observed motor recovery. When assessing whether motor recovery has occurred, several factors influence therapist decision-making process:

- **Muscle Group Tested:** The size and location of the muscle group play a crucial role. Smaller muscles, particularly those in the hand, are often easier to test accurately due to better positioning control and less influence from tester strength limitations.
- **Consistency and Fluctuations in Scores:** Fluctuating scores over time can indicate instability rather than genuine recovery. For instance, oscillations between grade 4+ and grade 5 may suggest measurement errors (inter-rater or intra-rater) rather than actual motor improvement. When interpreting MMT scores, several critical factors should be considered to ensure accuracy and reliability. These factors include the motor score recorded at baseline, which provides a benchmark for assessing changes over time. The direction and magnitude of change from baseline are crucial indicators of treatment effectiveness, whether positive or negative. Additionally, the duration over which a consistent score persists offers insights into the stability of motor improvements. It’s essential to recognize that the scale of change in MMT scores may not be uniform across all levels, as the scale is nominal. For instance, moving from a score of 1 to 2 does not necessarily represent the same functional change as progressing from 4 to 5. Therefore, the clinical significance of each change must be carefully evaluated within the context of the scale’s limitations. Furthermore, the overall arc of change, encompassing the trajectory of motor improvements over time, provides a comprehensive view of treatment outcomes.
- **Trend in MMT Scores:** An increase followed by a significant decrease in MMT scores over time would be interpreted as a lack of sustained recovery.
- **Confidence in Assessment:** In cases where the evidence for motor recovery based on MMT scores is inconclusive, the therapist adopt a conservative approach and lean towards labeling it as “no recovery” for those muscle groups.
- **Nominal Scale Considerations:** Recognizing that the MMT scale is nominal, changes between grades do not equate to equal increments in motor function. Each grade change (e.g., from 1 to 2, 2 to 3) represents a different level of functional improvement.
- **Baseline Motor Score and Change:** The initial motor score and the magnitude of change from baseline provide critical context for determining whether meaningful recovery has occurred. Moreover, it’s important to note that motor recovery can occur independent of direct FEST effects, such as when improvements are observed within a timeframe shorter than the typical treatment duration of 15 days.
- **Duration of Improvement:** Time is a crucial factor. For changes to be considered meaningful, improvements should persist for at least 20 days.
- **Spasticity:** Botox injections are aimed at managing complications or enhancing function but can temporarily weaken specific muscles, impacting assessment outcomes at certain time points. Noting the use of medications like Botulinum Toxin Type A (BoNTA) or oral medications to manage hypertonicity is essential, as these can significantly affect muscle movements. Medication peaks and troughs in the bloodstream can influence the smoothness or jerkiness of movements observed.
- **Support systems and resources outside of therapy sessions:** such as family support for home exercises or access to community-based therapies, may influence motor recovery outcomes positively or negatively.
- **Duration and frequency of FES interventions:** Some individuals may not have received sufficient total intervention time or sessions to demonstrate noticeable motor improvements. The timing of assessments and FES sessions throughout the day can also affect results, as patients may exhibit varying strength levels depending on the time of day.
- **Fatigue:** Inconsistent MMT scoring may arise due to patient fatigue during assessments, influenced by prior therapy sessions, daily activities, or health conditions like infections.
- **Reliability:** Consideration of interrater reliability in MMT assessments ensures consistency and validity in score interpretation across different evaluators. For example, interrater reliability for certain muscles tested, such as the 1st dorsal interossei or flexor digitorum superficialis, may not be well-validated in the SCI population, potentially contributing to variability in scoring.
- **Patient survey:** To enhance assessment comprehensiveness, implementing measures for functional changes and incorporating qualitative feedback through patient interviews or surveys can provide valuable insights into the holistic impact of FES. Finally, establishing clinical significance thresholds for MMT scores specific to each muscle tested would further refine the interpretation of motor recovery outcomes in response to FEST interventions.
- **Compensatory movement:** Controlling for compensatory movements is equally important to obtain accurate data. During assessments, preventing trunk or shoulder compensations is crucial, as these can distort the readings and affect the interpretation of muscle responses. Compensatory movements, often employed by individuals with SCI to overcome gravity, can inadvertently activate muscles at elbow, shoulder, and wrist joints, complicating the assessment of targeted muscles.
- **Positioning devices:** Consistency in the use of positioning devices for comfort and limb access is vital for maintaining assessment integrity. Sessions can be lengthy, and using consistent methods such as lap trays or pillows to support limbs ensures comfort and standardizes conditions across assessors. This consistency helps mitigate discomfort for participants and ensures reliable assessments of muscle response to FEST interventions.
- **Sensitivity of the grading scale:** In recent observations, a consistent timeframe of 40 to 60 days appears to be pivotal for observing motor recovery onset. Notably, there have been fluctuations observed within the 0-2 grading range. Specifically, in muscles initially graded as 0, we are increasingly observing the emergence of motor activity following intensive neuromodulation techniques. This trend underscores the necessity for a more nuanced 0-2 grading scale to accurately capture these developments. Muscles progressing from grade 0 to grade 1 are becoming more common, yet many do not advance to grade 2, highlighting a significant gap in current grading scales. Historically, the focus has been on grade 3 as a functional milestone, with changes below this level often deemed less relevant for functional outcomes. However, with advancements in neuromodulation showing potential for previously inactive muscles to exhibit some activity, there is now a critical need for clinical grading systems capable of capturing these subtle but meaningful changes. As we navigate this era of expanding therapeutic possibilities, a refined grading system will be essential for accurately assessing and documenting the evolving motor capabilities of individuals undergoing neuromodulatory treatments. We adopted the following approach. If a muscle is initially graded as a 1 or 2, re-evaluate and assign a grade based on the following additional definitions:

`o` Grade 1: Indicates a flicker with no observable movement of the joint.
`o` Grade 2-: Represents movement that is 50% or less of the full range in an anti-gravity position.
`o` Grade 2: Demonstrates movement that covers 50% or more of the full range in an anti-gravity position.
`o` Grade 2+: Indicates movement where part of the range is performed against gravity and part is performed with anti-gravity positioning.
`o` Grade 3: Represents full range of movement performed entirely against gravity.

These additional grading criteria refine the assessment of muscle strength within the scale, providing clearer distinctions that enhance the precision and reliability of strength evaluations for research and clinical purposes.

- **Motor Score Significance:** Even small changes in motor scores (e.g., 0.5 points) are deemed clinically important as they can potentially enhance the patient’s functional abilities.

These considerations collectively enhance the interpretation and utility of MMT scores in assessing motor function and treatment response; and ensure a comprehensive evaluation of motor recovery outcomes, taking into account the specific characteristics and nuances of the muscles being assessed and the limitations of the assessment scale. Baseline motor function and ASIA level diagnosis are also essential factors influencing the potential for motor recovery. These criteria ensure that the evaluation process is rigorous and sensitive to both short-term and sustained improvements in motor function, thereby informing decisions on treatment efficacy and patient outcomes.

**Supplementary Table 1.**
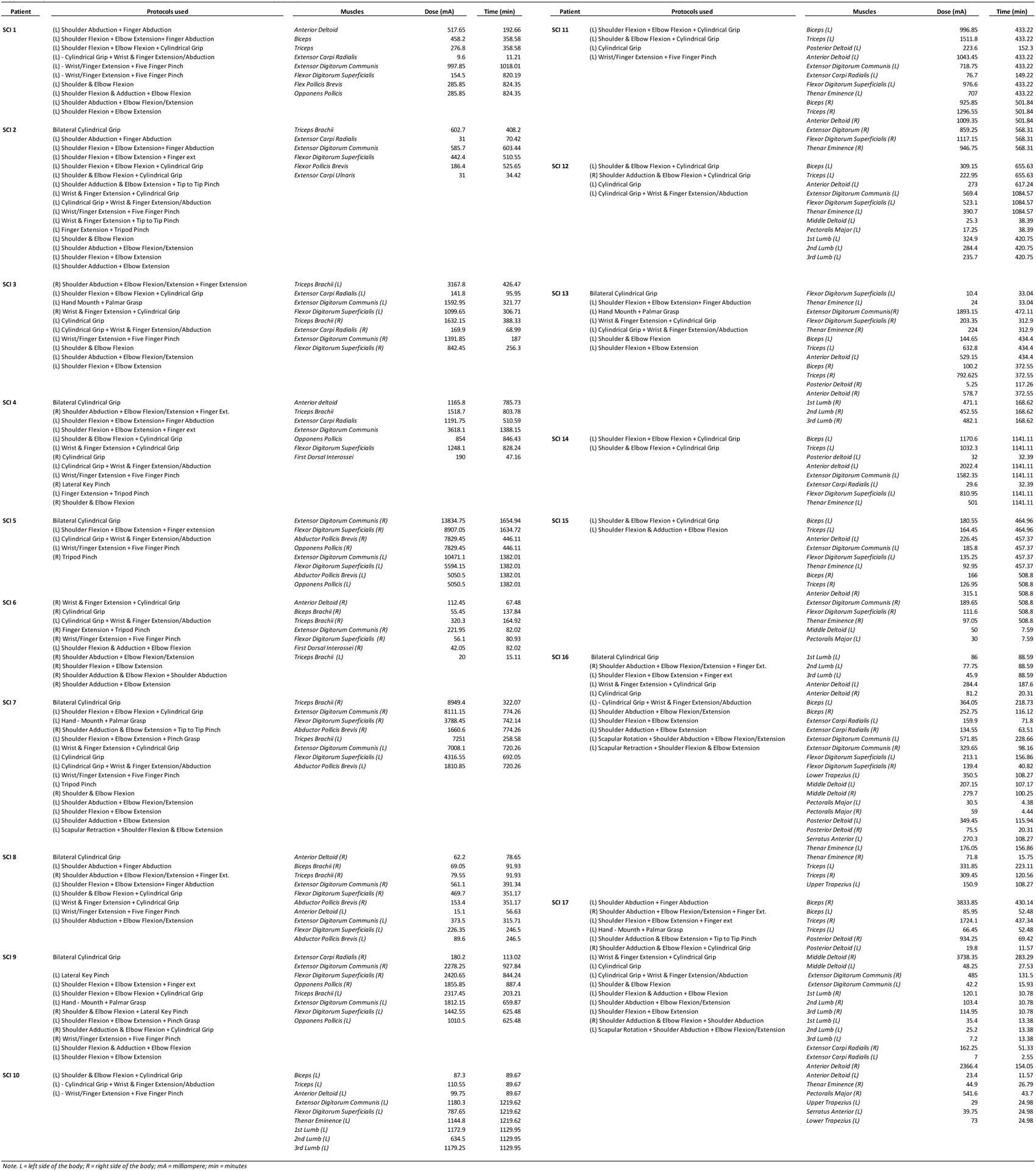

## References

1. Devivo, M. J. Epidemiology of traumatic spinal cord injury: Trends and future implications. Spinal Cord 50, 365–372 (2012).

2. Anderson, K. D. Targeting recovery: Priorities of the spinal cord-injured population. Journal of Neurotrauma 21, 1371–1383 (2004).

3. Lo, C., Tran, Y., Anderson, K., Craig, A. & Middleton, J. Functional Priorities in Persons with Spinal Cord Injury: Using Discrete Choice Experiments To Determine Preferences. Journal of Neurotrauma 33, 1958–1968 (2016).

4. Taccola, G., Sayenko, D., Gad, P., Gerasimenko, Y. & Edgerton, V. R. And yet it moves: Recovery of volitional control after spinal cord injury. Progress in Neurobiology 160, 64–81 (2018).

5. Moritz, C. et al. Non-invasive spinal cord electrical stimulation for arm and hand function in chronic tetraplegia: a safety and efficacy trial. Nat Med 30, 1276–1283 (2024).

6. Singer, B. Functional Electrical Stimulation of the Extremities in the Neurological Patient: A Review. Australian Journal of Physiotherapy 33, 33–42 (1987).

7. Marquez-Chin, C. & Popovic, M. R. Functional electrical stimulation therapy for restoration of motor function after spinal cord injury and stroke: A review. BioMedical Engineering Online 19, 1–25 (2020).

8. Kapadia, N., Moineau, B. & Popovic, M. R. Functional Electrical Stimulation Therapy for Retraining Reaching and Grasping After Spinal Cord Injury and Stroke. Frontiers in Neuroscience 14, (2020).

9. Velstra, I. M., Bolliger, M., Krebs, J., Rietman, J. S. & Curt, A. Predictive value of upper limb muscles and grasp patterns on functional outcome in cervical spinal cord injury. Neurorehabilitation and Neural Repair 30, 295–306 (2016).

10. Kirshblum, S., Snider, B., Eren, F. & Guest, J. Characterizing Natural Recovery after Traumatic Spinal Cord Injury. Journal of Neurotrauma 18, 1–18 (2021).

11. Kirshblum, S. C. & O’Connor, K. C. Predicting neurologic recovery in traumatic cervical spinal cord injury. Archives of Physical Medicine and Rehabilitation 79, 1456–1466 (1998).

12. Balbinot, G. et al. Segmental motor recovery after cervical spinal cord injury relates to density and integrity of corticospinal tract projections. Nature Communications 14, 723 (2023).

13. Sangari, S. & Perez, M. A. Prevalence of spasticity in humans with spinal cord injury with different injury severity. Journal of Neurophysiology 128, 470–479 (2022).

14. Sangari, S., Lundell, H., Kirshblum, S. & Perez, M. A. Residual descending motor pathways influence spasticity after spinal cord injury. Annals of Neurology 86, 28–41 (2019).

15. Sangari, S., Kirshblum, S., Guest, J. D., Oudega, M. & Perez, M. A. Distinct patterns of spasticity and corticospinal connectivity following complete spinal cord injury. The Journal of physiology 599, 4441–4454 (2021).

16. Anderson, K. D. et al. Multicentre, single-blind randomised controlled trial comparing MyndMove neuromodulation therapy with conventional therapy in traumatic spinal cord injury: a protocol study. BMJ open 10, e039650 (2020).

17. Committee, A. and Isc. I. S. The 2019 revision of the International Standards for Neurological Classification of Spinal Cord Injury (ISNCSCI)—What’s new? Spinal Cord 57, 815–817 (2019).

18. Ciesla, N., et al. Manual Muscle Testing: A Method of Measuring Extremity Muscle Strength Applied to Critically Ill Patients. Journal of Visualized Experiments : JoVE (2011) doi:10.3791/2632.

19. Schmitt, W. H. & Cuthbert, S. C. Common errors and clinical guidelines for manual muscle testing: ‘the arm test’ and other inaccurate procedures. Chiropractic & Osteopathy 16, 16 (2008).

20. Hislop, H. J. & Montgomery, J. Daniels and Worthingham’s Muscle Testing: Techniques of Manual Examination. (WB Saunders, Philadelphia, 1995).

21. Kirshblum, S. C. et al. Reference for the 2011 revision of the International Standards for Neurological Classification of Spinal Cord Injury. Journal of Spinal Cord Medicine 34, 547–554 (2011).

22. Kalsi-Ryan, S., Curt, A., Fehlings, M. G. & Verrier, M. C. Assessment of the hand in tetraplegia using the Graded Redefined Assessment of Strength, Sensibility and Prehension (GRASSP): Impairment versus function. Topics in Spinal Cord Injury Rehabilitation 14, 34–46 (2009).

23. Anderson, K. D. et al. Multi-center, single-blind randomized controlled trial comparing functional electrical stimulation therapy to conventional therapy in incomplete tetraplegia. Front. Rehabilit. Sci. 3, 995244 (2022).

24. Hermens, H. J., Freriks, B., Disselhorst-Klug, C., Rau, G. & Ray, G. Development of recommendations for SEMG sensors and sensor placement procedures. Journal of Electromyography and Kinesiology 10, 361–374 (2000).

25. Perotto, A. Anatomical Guide for the Electromyographer: The Limbs and Trunk. (Charles C Thomas, 2011).

26. Thongpanja, S., Phinyomark, A., Phukpattaranont, P. & Limsakul, C. Mean and Median Frequency of EMG Signal to Determine Muscle Force based on Time-Dependent Power Spectrum. Electronics and Electrical Engineering 19, (2013).

27. Thomas, C. K., Dididze, M., Martinez, A. & Morris, R. W. Identification and classification of involuntary leg muscle contractions in electromyographic records from individuals with spinal cord injury. Journal of Electromyography & Kinesiology 24, 747–754 (2014).

28. Zijdewind, I., Bakels, R. & Thomas, C. K. Motor unit firing rates during spasms in thenar muscles of spinal cord injured subjects. Frontiers in Human Neuroscience 8, 922 (2014).

29. Zijdewind, I. & Thomas, C. K. Motor unit firing during and after voluntary contractions of human thenar muscles weakened by spinal cord injury. Journal of Neurophysiology 89, 2065–2071 (2003).

30. Zijdewind, I. & Thomas, C. K. Spontaneous motor unit behavior in human thenar muscles after spinal cord injury. Muscle & Nerve 24, 952–962 (2001).

31. Zijdewind, I. & Thomas, C. K. Firing patterns of spontaneously active motor units in spinal cord-injured subjects. Journal of Physiology 590, 1683–1697 (2012).

32. Maffiuletti, N. A., Roig, M., Karatzanos, E. & Nanas, S. Neuromuscular electrical stimulation for preventing skeletal-muscle weakness and wasting in critically ill patients: a systematic review. BMC Medicine 11, 137 (2013).

33. Calancie, B., Molano, R., Broton, J. G., Bean, J. A. & Alexeeva, N. RELATIONSHIP BETWEEN EMG AND MUSCLE FORCE AFTER SPINAL CORD INJURY. J Spinal Cord Med 24, 19–25 (2000).

34. Popovic, M. R. et al. Functional Electrical Stimulation Therapy of Voluntary Grasping Versus Only Conventional Rehabilitation for Patients With Subacute Incomplete Tetraplegia: A Randomized Clinical Trial. Neurorehabilitation and Neural Repair 25, 433–442 (2011).

35. Ditunno, J. F., Cohen, M. E., Hauck, W. W., Jackson, A. B. & Sipski, M. L. Recovery of upper-extremity strength in complete and incomplete tetraplegia: A multicenter study. Archives of Physical Medicine and Rehabilitation 81, 389–393 (2000).

36. Mange, K., Ditunno, J., Herbison, G., & Jaweed, MM. Recovery of strength at the zone of injury in motor complete and motor incomplete cervical spinal cord injured patients. Arch Phys Med Rehabil 71, 562–565 (1990).

37. Waters, R. L. Motor and sensory recovery following incomplete tetraplegia. Arch Phys Med Rehabil 75, 306–311 (1994).

38. Khorasanizadeh, M. H. et al. Neurological recovery following traumatic spinal cord injury: A systematic review and meta-analysis. Journal of Neurosurgery: Spine 30, 683–699 (2019).

39. McDonald, J. W., Becker, D., Sadowsky, C. L., Conturo, T. E. & Schultz, L. M. Late recovery following spinal cord injury. Journal of Neurosurgery 97, 405–406 (2002).

40. Kirshblum, S., Millis, S., McKinley, W. & Tulsky, D. Late neurologic recovery after traumatic spinal cord injury. Archives of Physical Medicine and Rehabilitation 85, 1811–1817 (2004).

41. Lu, D. C. et al. Engaging Cervical Spinal Cord Networks to Reenable Volitional Control of Hand Function in Tetraplegic Patients. Neurorehabilitation and Neural Repair 30, 951–962 (2016).

42. Snoek, G. J., Ijzerman, M. J., Hermens, H. J., Maxwell, D. & Biering-Sorensen, F. Survey of the needs of patients with spinal cord injury: Impact and priority for improvement in hand function in tetraplegics. Spinal Cord 42, 526–532 (2004).

43. Fawcett, J. W. et al. Guidelines for the conduct of clinical trials for spinal cord injury as developed by the ICCP panel: Spontaneous recovery after spinal cord injury and statistical power needed for therapeutic clinical trials. Spinal Cord 45, 190–205 (2007).

44. Hupp, M., Pavese, C., Bachmann, L. M., Koller, R. & Schubert, M. Electrophysiological Multimodal Assessments Improve Outcome Prediction in Traumatic Cervical Spinal Cord Injury. Journal of Neurotrauma 35, 2916–2923 (2018).

45. Kramer, J. L. K., Lammertse, D. P., Schubert, M., Curt, A. & Steeves, J. D. Relationship between motor recovery and independence after sensorimotor-complete cervical spinal cord injury. Neurorehabilitation and Neural Repair 26, 1064–1071 (2012).

46. Steeves, J. D. et al. Extent of spontaneous motor recovery after traumatic cervical sensorimotor complete spinal cord injury. Spinal Cord 49, 257–265 (2011).

47. Wilson, J. R., Cadotte, D. W. & Fehlings, M. G. Clinical predictors of neurological outcome, functional status, and survival after traumatic spinal cord injury: a systematic review. Journal of neurosurgery. Spine 17, 11–26 (2012).

48. Brüningk, S. C. et al. Prediction of segmental motor outcomes in traumatic spinal cord injury: Advances beyond sum scores. Experimental Neurology 380, 114905 (2024).

49. Håkansson, S. et al. Data-driven prediction of spinal cord injury recovery: An exploration of current status and future perspectives. Experimental Neurology 380, 114913 (2024).

50. Ahuja, C. S. et al. Traumatic spinal cord injury. Nature Reviews Disease Primers 3, (2017).

51. Morecraft, R. J. et al. Terminal distribution of the corticospinal projection from the hand/arm region of the primary motor cortex to the cervical enlargement in rhesus monkey. The Journal of comparative neurology 521, 4205–35 (2013).

52. Lemon, R. N. & Morecraft, R. J. The evidence against somatotopic organization of function in the primate corticospinal tract. Brain (2022) doi:10.1093/brain/awac496.

53. Rosenzweig, E. S. et al. Extensive spontaneous plasticity of corticospinal projections after primate spinal cord injury. Nature Neuroscience 13, 1505–1512 (2010).

54. Jaja, B. N. R. et al. Trajectory-Based Classification of Recovery in Sensorimotor Complete Traumatic Cervical Spinal Cord Injury. Neurology 96, e2736–e2748 (2021).

55. Berger, M. J. et al. Segmental infralesional pathological spontaneous activity in subacute traumatic spinal cord injury. Muscle and Nerve 69, 403–408 (2024).

56. Zijdewind, I., Gant, K., Bakels, R. & Thomas, C. K. Do additional inputs change maximal voluntary motor unit firing rates after spinal cord injury? Neurorehabilitation & Neural Repair 26, 58–67 (2012).

57. Bryden, A., Kilgore, K. L. & Nemunaitis, G. A. Advanced assessment of the upper limb in tetraplegia: A three-tiered approach to characterizing paralysis. Topics in Spinal Cord Injury Rehabilitation 24, 206–216 (2018).

58. Bryden, A. M. et al. Upper extremity assessment in tetraplegia: The importance of differentiating between upper and lower motor neuron paralysis. Archives of Physical Medicine and Rehabilitation 97, S97–S104 (2016).

59. Balbinot, G. et al. Properties of the surface electromyogram following traumatic spinal cord injury: a scoping review. Journal of NeuroEngineering and Rehabilitation 18, 105 (2021).

60. Balbinot, G. et al. The use of surface EMG in neurorehabilitation following traumatic spinal cord injury: a scoping review. Clinical Neurophysiology (2022) doi:10.1016/j.clinph.2022.02.028.

61. Le-Rademacher, J., Gunn, H., Yao, X. & Schaid, D. J. Clinical Trials Overview: From Explanatory to Pragmatic Clinical Trials. Mayo Clinic Proceedings 98, 1241–1253 (2023).

62. Fehlings, M., Singh, A., Tetreault, L., Kalsi-Ryan, S. & Nouri, A. Global prevalence and incidence of traumatic spinal cord injury. CLEP 309 (2014) doi:10.2147/CLEP.S68889.

63. Craven, B. C. & Morris, A. R. Modified Ashworth scale reliability for measurement of lower extremity spasticity among patients with SCI. Spinal Cord 48, 207–213 (2010).

